# A mixed-methods study of the scale-up and delivery of Seasonal Malaria Chemoprevention in pastoralist communities of northwest Kenya

**DOI:** 10.64898/2026.02.10.26346051

**Authors:** Diana Menya, Emmah Kimachas, Beatrice Rotich, Catherine Kafu, Joseph Kipkoech, Lucy Abel, Rebecca Lokwang, Mireya Dorado, David Ekai, Suzanne Van Hulle, Akinola Shonde, Victor Osiare, Edwin Mbugua, Wendy Prudhomme O’Meara

## Abstract

Seasonal Malaria Chemoprevention (SMC) is a promising intervention for Turkana, Northern Kenya, where malaria transmission is highly seasonal. Traditional malaria control methods, such as indoor residual spraying (IRS) and insecticide-treated nets (ITNs), are impractical due to the population’s semi-nomadic lifestyle, temporary dwellings, sparse settlements, and limited access to health facilities. In 2024, following the WHO’s updated guidance on SMC use, this intervention was implemented in Turkana Central for the first time, involving five monthly cycles of sulphadoxine-pyrimethamine with amodiaquine (SPAQ). To assess the program’s feasibility, a mixed-methods study was conducted at the end of the campaign. Survey data from a randomly selected, representative sample of 449 households with 680 eligible children were analyzed using multi-level logistic regression to compare partial versus complete SMC adopters, accounting for clustering. It was supplemented by qualitative interviews involving 45 caregivers to explore barriers and facilitators to SMC adoption. The campaign achieved notable success, with 97% of children receiving at least one SMC cycle (95% CI: 94–99%), and 71% receiving all 5 cycles (95% CI: 66–75%), primarily through door-to-door delivery. The quality of delivery was evident, as 99% of caregivers reported direct observation of the first dose and proper instructions for subsequent days. Adherence to day 2 and 3 medication remained high at 95% (95% CI: 93.5–98.1). Regression analysis suggested that households familiar with their Community Health Promoter (CHP) and who communicated SMC information had lower odds of missing cycles. In contrast, children from wealthier families showed a 93% higher odds of missing cycles. Qualitative findings revealed that positive caregiver experiences with SMC effectiveness drove continuation, while late adoption was linked to illness/ineligibility, uncertainty, and rumors. Overall, these findings indicate that high and sustained SMC coverage is feasible in marginalized settings through adaptive delivery strategies and leveraging of trusted CHP networks, establishing a scalable model for similar mobile populations.

## INTRODUCTION

Malaria remains a significant public health problem in Kenya, where 75% of the population is at risk of the disease, including 13 million people in endemic areas and another 19 million in epidemic-prone and seasonal transmission areas. Malaria transmission and infection risk in Kenya are primarily determined by climatic and environmental factors such as altitude, rainfall patterns, and temperature, leading to considerable variation in malaria prevalence by season and across geographic zones [1].

Turkana County, located in northwestern Kenya, borders Uganda to the west, South Sudan to the north, and Ethiopia to the northeast, with Lake Turkana forming the county’s eastern border. Turkana is a vast region with a sparse population of 1.3 million, 13% of whom are under the age of five. Like other pastoral communities in Sub-Saharan Africa, the Turkana population is economically marginalized, often lacking access to the basic services available to more settled populations [2]. Historically, malaria transmission in this region was thought to occur only in isolated areas and had the potential to become epidemic following unusual rainfall events. However, a reactive case detection study conducted in Turkana Central revealed the presence of both symptomatic and asymptomatic *P. falciparum* infections throughout the year, confirming that malaria transmission is endemic with a prevalence of 33% among asymptomatic community members [3].

Malaria transmission in this region is highly seasonal. Approximately 80% of cases occur within 5 months, according to the Kenya Health Information System. However, mobile pastoralists are challenging to reach with conventional health delivery systems. In Turkana, over half of families seasonally migrate with their animals [4]. In addition, most dwellings are made from temporary materials, household members sleep outside, and families have poor access to formal health services, resulting in low penetration of appropriate malaria case management or vector control tools such as insecticide-treated nets. Due in part to these challenges, malaria transmission continues largely unchecked. A recent report [5] demonstrated that individuals who travel with the herds have a two-fold higher prevalence of malaria than those who remain in the home village, calling for intervention strategies amenable to mobile lifestyles.

Seasonal Malaria Chemoprevention (SMC) was identified as a promising strategy for Turkana [6], given the seasonality of malaria transmission [7], the large proportion of children under 5, and the prevention gap for these families. However, the mobility of the population and the remoteness of many villages make it challenging for an intervention that requires monthly contact with families throughout the high transmission season. Formative work was conducted before the beginning of the malaria transmission season in Turkana to inform the design of an SMC programme tailored to the community’s needs. Potential strategies were further informed by experience with previous mass-distribution campaigns, such as polio immunization and trachoma treatment. Additionally, healthcare workers and community leadership identified children most likely to be missed during the SMC campaign. The engagement of grassroots leadership structures and mobilization were highlighted as critical aspects to the success of SMC [7].

SMC is a promising intervention in Turkana, where population mobility and poor health infrastructure exacerbate malaria transmission and burden. This study uses a mixed-methods approach to evaluate the coverage and delivery of the adapted SMC program in Turkana, Kenya. We aim to determine whether high SMC coverage can be attained and maintained across multiple cycles in remote, often highly mobile populations. We describe the feasibility and community perceptions of SMC among caretakers who fully participated in SMC, those whose children did not receive all cycles, and those whose children did not receive SMC at all. Findings from this study inform how SMC can be deployed at a programmatic scale in similar regions experiencing intense seasonal malaria transmission.

## METHODS

### SMC campaign

The Turkana County Ministry of Health, with support from Catholic Relief Services (CRS), delivered SMC in Turkana Central from June 13, 2024, to October 7, 2024, incorporating 5 days of drug distribution at 28-day intervals [8]. Sulfadoxine-pyrimethamine (SP) and Amodiaquine (AQ) were given over 3 days: SP and AQ as directly observed therapy on day 1, with the child’s caregiver administering AQ on days 2 and 3. The programme targeted 38,585 children in one sub-county at the beginning of the malaria season. Community structures, livelihoods, migration patterns and malaria burden vary widely across the wards in Turkana Central subcounty and were expected to influence SMC acceptability and reach. This called for SMC delivery approaches to be tailored to context [7]. Drug distribution was primarily door-to-door, supplemented by fixed points and mobile fixed points in areas with vast distances between households. Integrated outreaches combined SMC with Antenatal Care Services (ANC), nutrition screening and referral, and malaria case management, targeting hard-to-reach communities. These additional services provided comprehensive healthcare and created demand for SMC among families living far from health facilities.

### Study area

This mixed-methods study was carried out in the SMC delivery sub-county, Turkana Central, which has 5 wards. There are two peri-urban wards, Township and Kanamkemer, where the population is more settled and clustered. The other three wards, Kalokol, Kangatosa and Kerio, are more rural, with sparse settlements and frequent population movement.

Coverage was measured through a quantitative survey, while the nuances in SMC perspectives, experiences and motivators that may have influenced SMC uptake were explored using a qualitative approach. The qualitative description sought to identify the key facilitators and barriers to full, partial, or non-adoption of SMC (those who received SMC in all months, those who received SMC in some months, categorised as either late adopters or dropouts, and those who did not receive any SMC at all).

### Sampling procedures for the quantitative survey

Households that participated in the cross-sectional survey were selected through a multistage sampling process from the ward to the village level. A complete list of villages (n=644) was stratified by ward, and 9 villages were randomly selected from each stratum with equal probability. Within each village, all households with children aged 6-59 months were listed from the Ministry of Health registers and supplemented by community leaders to ensure the lists were complete and included households not mapped into the health network. Ten households were then selected by systematic random sampling with a random starting point and an interval determined by the village size. Five replacement households were randomly selected a priori. Unavailable households were visited at a later date and only replaced if they were still away after a second visit. The target enrollment was 450 households (10 per village, in 9 villages in each of 5 wards) which was sufficient to ensure 87% power to measure a 15 percent point difference in coverage of the primary outcome (at least one cycle of SMC) between the poorest and least poor households based on a precision of 5% and a design effect of 1.2 to account for clustering of households in villages.

### Data Collection

Households with at least one child aged 6-59 months eligible for SMC in accordance with the WHO criteria were eligible to participate. Households whose primary residence was in Turkana Central but who migrated in and out of the sub-county during the SMC period were included. In contrast, households visiting for short periods were excluded. The primary caretaker of the child was interviewed using a structured tool to elicit information on household demographics, malaria prevention, recent illnesses, health-seeking behavior, health facility access, household socioeconomic indicators, travel during the SMC period and overall engagement in the SMC programme. Interviews were conducted in either Kiswahili or Nga’Turkana, according to the interviewee’s preference. Survey data were directly entered into tablets using the KOBOCollect Android app [9]. Household members with malaria symptoms on the data collection day were offered a malaria rapid diagnostic test (mRDT). A dried blood spot sample was collected from all household members by dropping 5-10 μl of capillary blood from a finger prick onto a filter paper and allowing it to dry completely.

### Study Outcomes

SMC coverage was measured as the proportion of households contacted during each of the 5 monthly SMC cycles. Those who did not complete all five cycles were further categorized into those who started after the first cycle (late adopters) and those who discontinued before the last cycle (dropouts or loss). Adherence to the SMC regimen was ascertained by calculating the proportion of children who completed all three doses of SMC drugs within the correct time window for the last cycle in which they participated. We also recorded information about adverse drug reactions. All information was based on the caregiver’s self-report, supplemented by documentation of cycles on the SMC card. Data were exported, then cleaned, curated, and key outcomes visualised in R (RStudio 2023) and STATA 17. All summary statistics were estimated using design weights, which accounted for villages within wards and households within villages. Regression analyses were used to understand factors associated with partial versus complete SMC for each observed child. Further exploratory analyses compared children with partial SMC to those who dropped out and partial versus late adopters. Multi-level logistic regression models included both household and child-level variables and accounted for clustering at the ward level via fixed effect and at the household level. Models that attempted to account for clustering at both the village and household levels failed to converge due to the paucity of mixed outcomes within households.

### Qualitative study participants

Participants of the qualitative survey were a subset of families that participated in the quantitative study and those identified through ’ground truth’ activities with community leaders. They were identified purposefully to fit the three categories that describe SMC adoption as described above (full SMC, no SMC and partial SMC) in equal representation across the wards. Additional consideration was made to ensure that families were selected from the peri-urban community, fisherfolk, pastoralists and farming communities in the riverine areas of Turkana Central. A total of 45 caregivers were selected from the three categories in equal proportions and were interviewed to elaborate on the challenges and successes of the delivery strategies adopted to reach eligible children.

### Qualitative data collection and analysis

Semi-structured in-depth interviews were conducted by trained research assistants in the language the participant was most comfortable with (English, Kiswahili, or Nga’ Turkana) and audio-recorded. The main domains explored using the interview guides were household health-seeking behaviours, malaria knowledge, malaria care-seeking behaviors, and perceptions of SMC over the course of the intervention (before SMC, during the SMC period, and at the end of the peak malaria season). Participants also gave recommendations for improving SMC delivery and uptake in future SMC rounds.

The audio-recorded interviews were transcribed and translated into English (where necessary). They were reviewed and exported to MAXQDA 24. Three qualitative codebooks were developed through a deductive approach based on the key domains identified in the three interview guides for full adopters, partial adopters and non-adopters. Within these main domains, the research team inductively identified sub-themes that emerged from the participants’ responses. Findings from both surveys were compared to explain coverage, identify differences in coverage, and identify the determinants most important in driving SMC participation in this nomadic community.

### Ethical considerations

The study protocol was reviewed and approved by the Moi University Institutional Research Ethics Committee (IREC) (Approval number FAN:0004733) and the Duke University Institutional Review Board. Written informed consent was obtained from a caregiver for household and SMC information. Written informed consent was also obtained before blood samples were collected from participants aged 18 years and older. Written informed consent from a parent or guardian was required for participants <18 years, along with assent for those aged 10 years or older. Written consent was also obtained from all participants of the qualitative survey.

## RESULTS

### Characteristics of the study population

Household interviews were conducted between October 9 and November 1, 2024. Data were collected from 449 households, 50 households in each of the five wards, except for Kerio Ward, where one sampled village had only nine eligible households (Table 1). The median household size was six members (IQR: 4–8), with a median of 2 SMC-eligible children per household (IQR: 1–2, maximum 6). Caregivers had a mean age of 33 years (SD: 10), and the majority were female (96%). Notably, 70% of caregivers had no formal education. The average travel time to the nearest health facility was 30 minutes, though it ranged from 2 minutes to 6 hours. Among the 733 SMC-eligible children identified in the sampled households, the majority were male (53%), with a mean age of 33 months (SD: 18). Here, we analyzed data from 680 children who had reached three months of age before the first cycle of SMC and were therefore eligible for all five cycles.

**Table 1.** Descriptive characteristics of sampled households.

| Variable | N | Households = 449' |
| --- | --- | --- |
| Household characteristics |  |  |
| Household size | 449 | 6.00 (4.00, 8.00) |
| Children under 5 | 448 |  |
| 1 |  | 187 (42%) |
| 2 |  | 215 (48%) |
| 3 |  | 39 (8.7%) |
| More than 3 |  | 7 (1.6%) |
| Time to facility | 449 | 30 (15, 60) |
| Own livestock | 449 | 262 (58%) |
| Travel with livestock | 262 | 16 (6.1%) |
| Travel with child under five | 40 | 17 (43%) |
| Income | 449 |  |
| Charcoal/Firewood |  | 96 (21%) |
| Farming/Agriculture |  | 19 (4.2%) |
| Fishing |  | 34 (7.6%) |
| Formal employment |  | 20 (4.5%) |
| Livestock-based |  | 42 (9.4%) |
| Other |  | 90 (20%) |
| Small business/Trade |  | 148 (33%) |

| Variable | N | Households = 449<sup>i</sup> |
| --- | --- | --- |
| Respondent characteristics |  |  |
| Respondent age | 458 | 30 (24, 38) |
| Respondent gender | 458 |  |
| Female |  | 438 (96%) |
| Male |  | 20 (4.4%) |
| Respondent education level | 142 |  |
| Completed primary |  | 16 (11%) |
| Completed Secondary |  | 33 (23%) |
| Higher/Tertiary |  | 15 (11%) |
| Some primary |  | 60 (42%) |
| Some Secondary |  | 18 (13%) |

<sup>i</sup>Median (Q1, Q3); n (%)

### Delivery of SMC

Ninety per cent (90%) of children who received SMC did so at home, where the SPAQ was delivered by a drug distributor (most often a community health promoter, CHP). Although few children received SMC outside their homes, in one sampled village, 85% received SMC at a fixed delivery point implemented in the third cycle. The survey assessed the quality of SMC delivery by the drug distributor and the caregiver’s adherence to the day 2 and day 3 SMC doses at each cycle. More than 99% of caregivers reported that drug distributors directly observed the first dose (Figure 1), 93.5% explained how to administer subsequent doses, and 98.9% reported that the drug distributor explained the types of side effects they might expect. Only 81% of caregivers had an SMC card. Ninety-five per cent (95% CI: 93.5-98.1) of caregivers reported administering day 2 and day 3 medications on time as directed.

**Figure 1:**
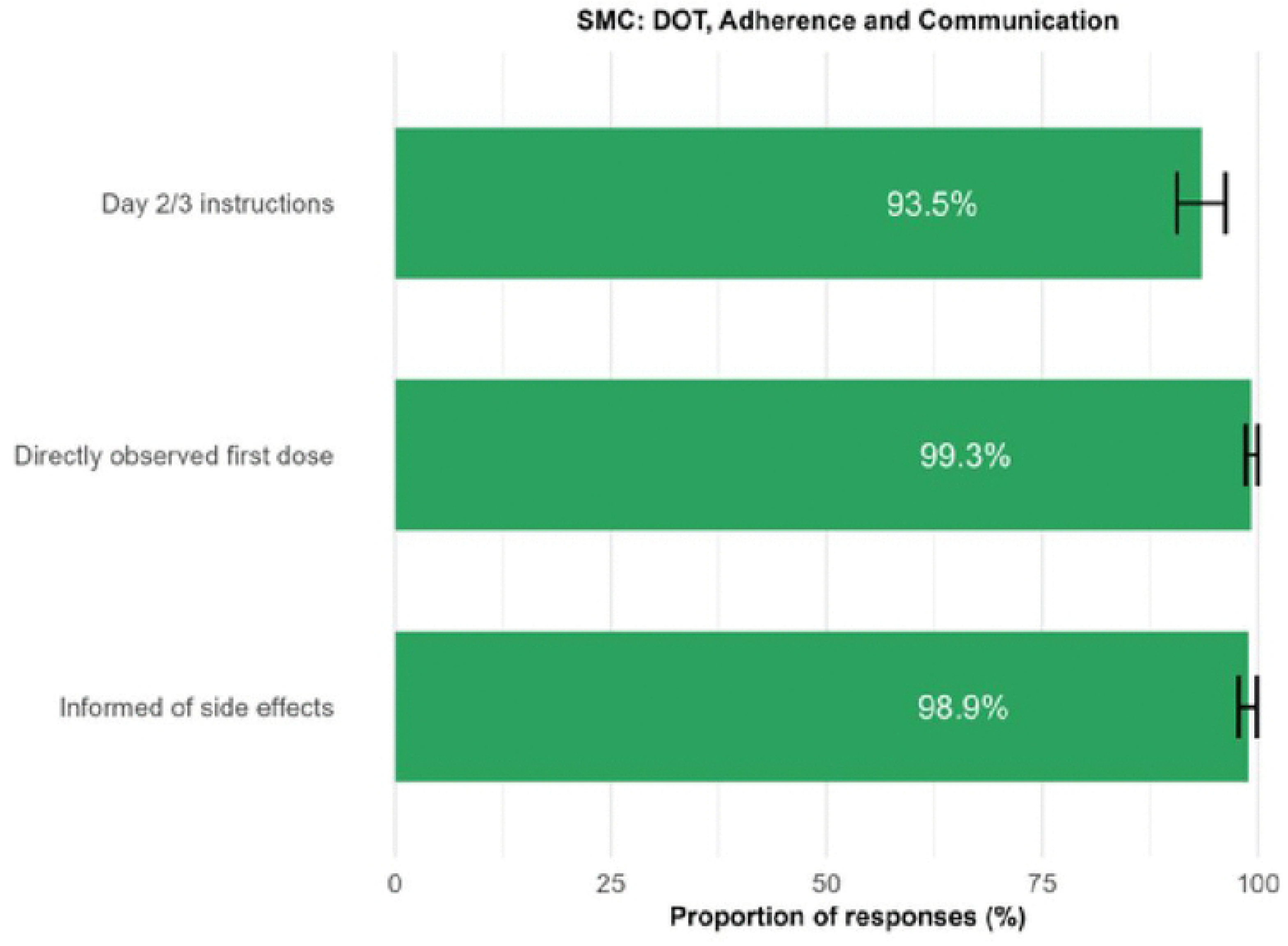
Proportion of households reporting directly observed first dose of SMC, receiving instructions from the drug distributor, and receiving information on possible side effects after SMC.

**Figure 2:**
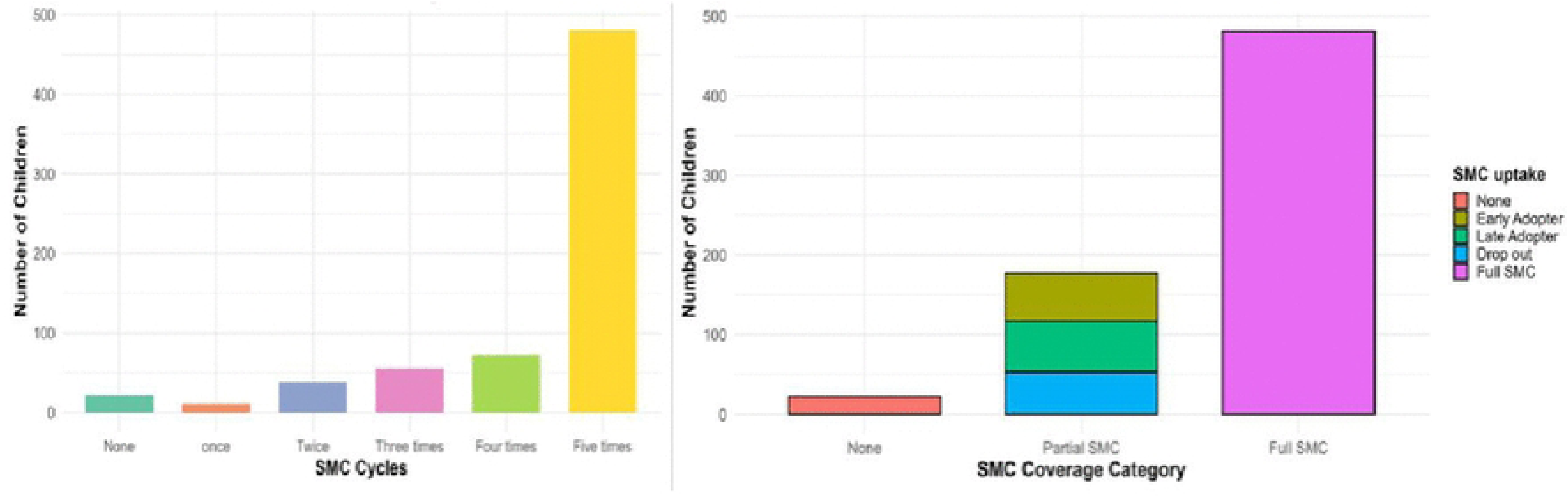
Number of children receiving 0-5 cycles of SMC (left) and the children receiving 1-4 cycles categorized as either late adopter, dropout or other partial (right)

instructions from the drug distributor about how to administer the second and third doses, and receiving information on possible side effects after SMC.

The quality of this interaction at the household level was important for caregivers’ confidence in SMC and their perceived self-efficacy in administering subsequent doses. Caregivers perceived the SMC distributors as being knowledgeable about malaria and SMC based on the SMC distributor’s profession as either a clinician or a CHP, coupled with the SMC training that they had received. Their mastery of SMC was illustrated through their provision of comprehensive information on SMC, demonstration of SMC administration and the actual administration of SMC. Partial adopters further revealed that being well informed of the potential side effects of SMC gave them confidence to administer the day 2 and 3 doses. Notably, this confidence in being able to give the day 2 and 3 doses was cited as a factor in continuing SMC in future cycles, and caregivers directly attributed this to the clear instructions and demonstration provided by the drug distributor.

Yes, he was well conversant, I understood the procedure of administering the dose from him; he was precise and clear. (SMC_PA_05)
**Interviewer:** Could you please narrate the entire process of administering SMC that you did on your own.
**Respondent:** It was easy. I put water in a cup, the exact amount the CHP had said. I then put the tablet in the water, it dissolved, then I gave the solution to the child.
**Interviewer:** What made it easier for you?
**Respondent:** My experience with administering medicine, plus the instructions that the CHP gave were simple and clear (SMC_PA_10)
I gave the [SMC] to the child that evening. I did what the CHP had told me. I did not panic because I had been informed of the side effects. (SMC_PA_02)

### Coverage of SMC

The SMC intervention was implemented over 5 cycles, conducted at 28-day intervals from June 13, 2024, to October 7, 2024. Nearly all eligible children received at least one cycle of SMC. Among children eligible to receive SMC from the first cycle, nearly all received at least one cycle (97%, 95% CI: 94-99%), and 71% (95% CI: 66–75%) completed all 5 cycles of SMC. Approximately 27% (95% CI: 22–30%) of children received partial SMC, defined as 1-4 cycles. Of the partial adopters, 31.8% (57/179) were ’late adopters’ who joined SMC after the first cycle, and 29.1% (52/179) dropped out before the final cycle.

### Factors correlated with SMC coverage

We hypothesised that key household characteristics could influence SMC coverage, especially household wealth, distance to a health facility, and community health network membership.

Households whose children completed all five cycles of SMC were markedly closer to a health facility than those who joined SMC late.

In contrast, children who dropped out of SMC were much closer to a facility compared to those who completed SMC or joined SMC late. A lower proportion of children who missed SMC completely or joined late knew their local CHP. Households in the ‘least poor’ category had more children with partial SMC (37%) than those in the middle (27%) and poorest (22.2%) wealth categories (Figure 3).

**Figure 3:**
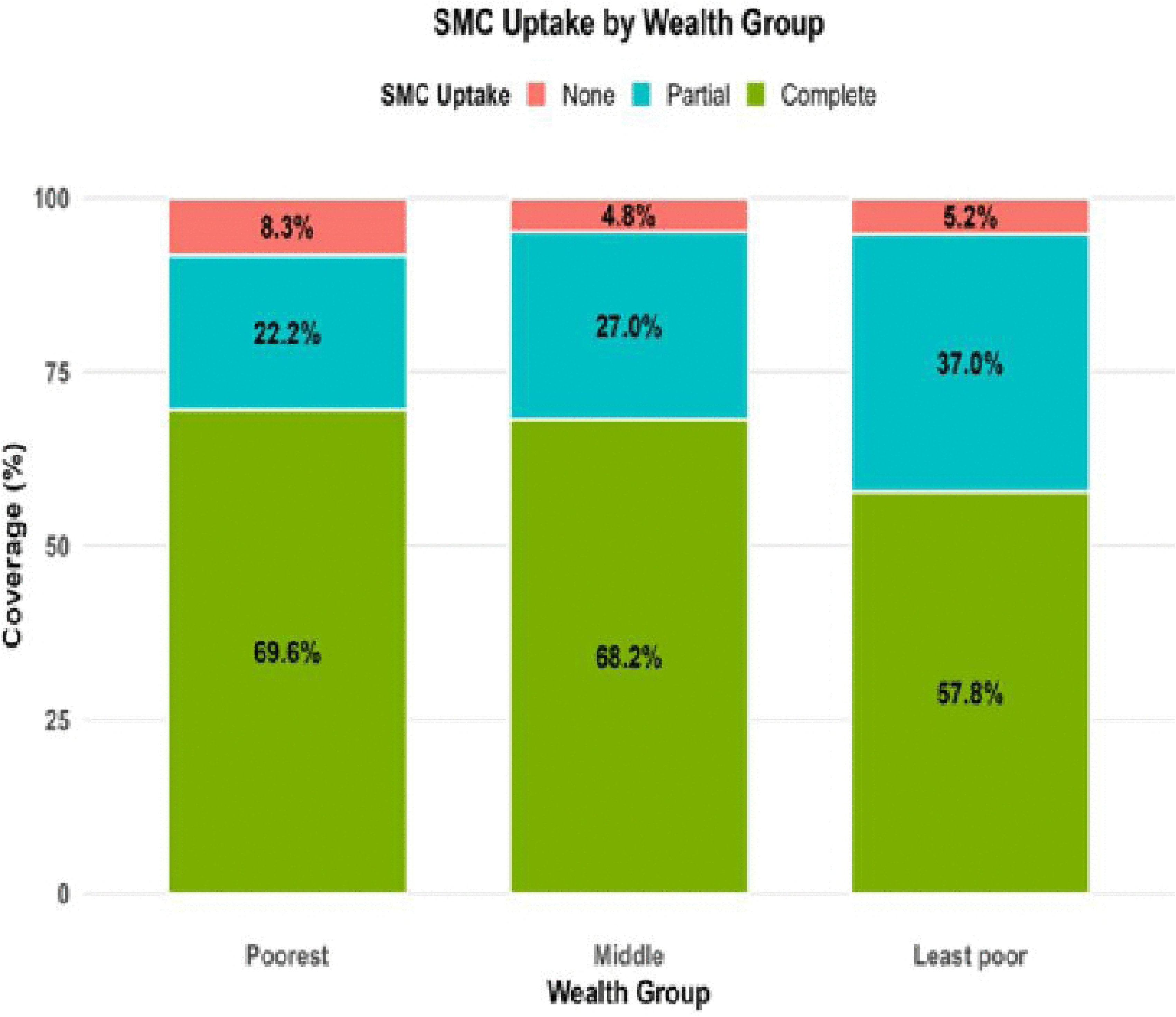
Full, partial and no SMC by wealth category.

We compared household and child characteristics between those with partial SMC and those with full SMC using a mixed-effects multivariable logistic regression (Table 2).

**Table 2:**
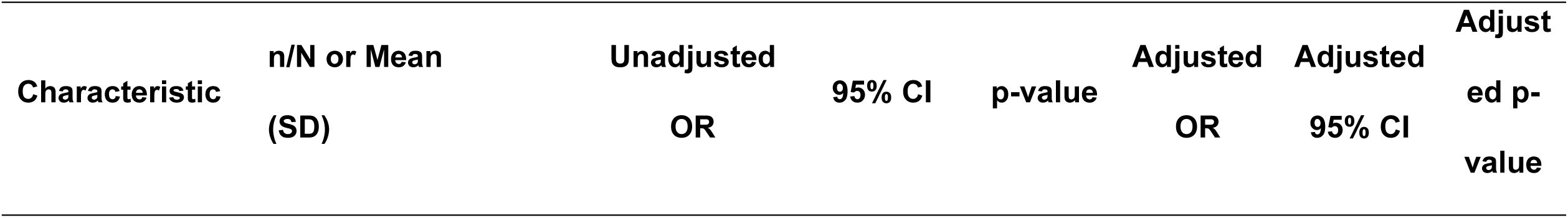
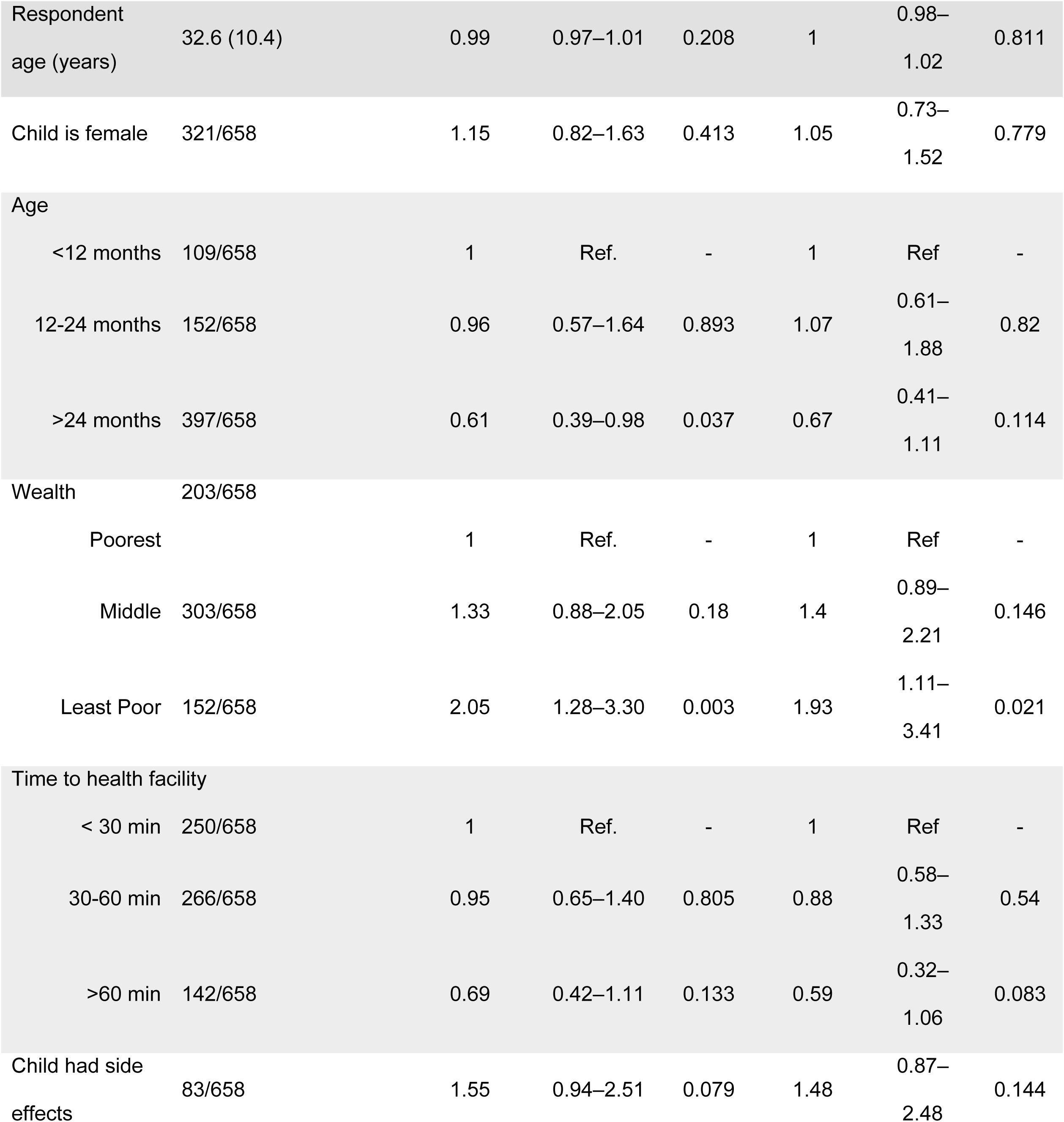
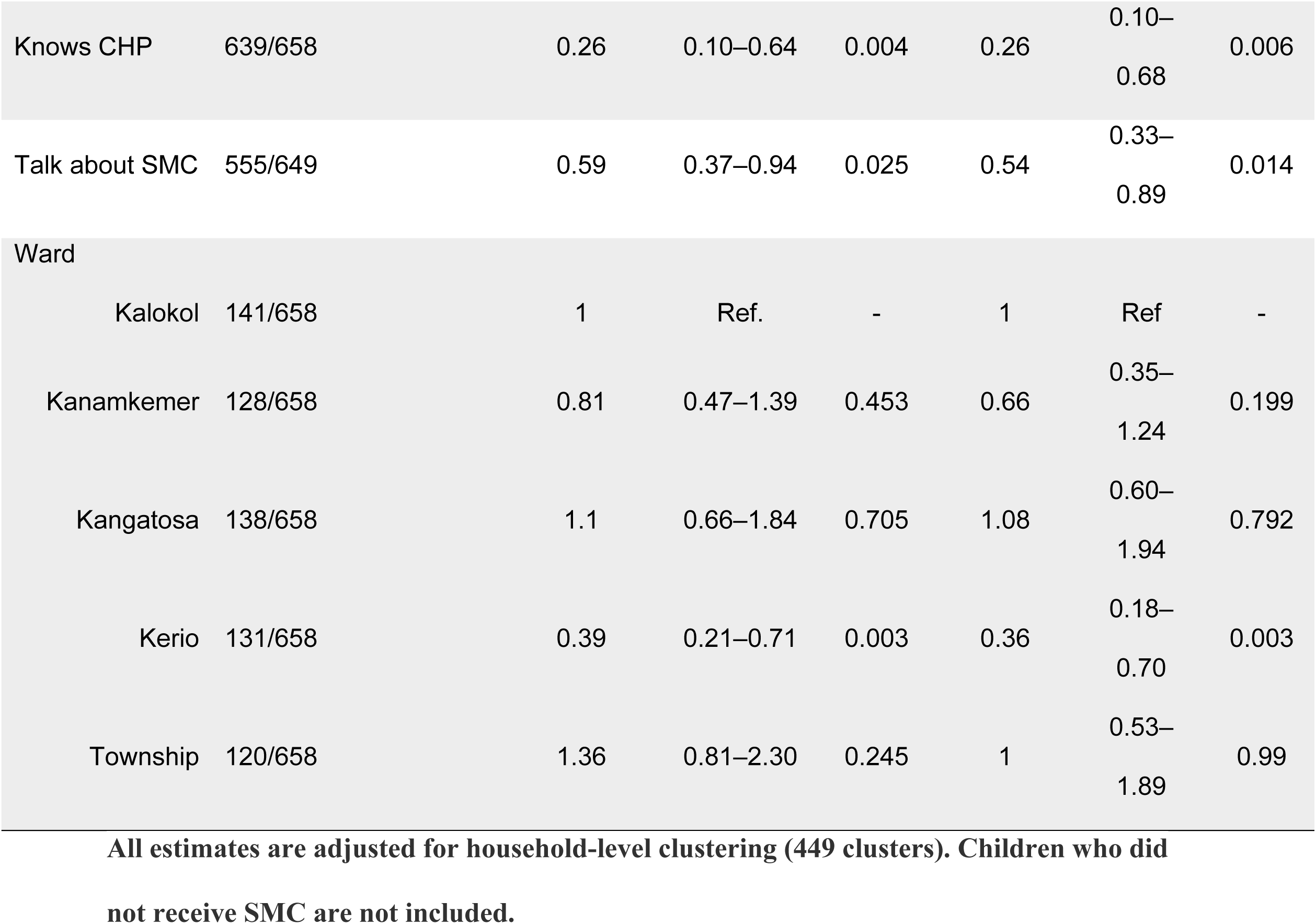
Logistic regression of factors related to partial versus complete SMC uptake in Turkana Central.

### All estimates are adjusted for household-level clustering (449 clusters). Children who did not receive SMC are not included

Caretakers who knew the CHP in their village had 74% lower odds of missing any cycles (Adj. OR=0.26, 95%CI:0.10-0.68). Similarly, talking about SMC with family or neighbors also significantly reduced the odds of missing any cycles (Adj OR=0.54, 95%CI:0.33-0.89). Those who lived furthest from the facility had lower odds of missing a cycle (Adj OR=0.57, 95%CI:0.31-1.01), and older children were also less likely to miss any cycles (Adj OR=0.67, 95%CI: 0.41-1.11). However, these observations fell slightly short of the criteria for statistical significance in the adjusted models. A child who experienced side effects was not less likely to complete all five cycles, but children from the least poor families had nearly double the odds of missing a cycle (Adj. OR=1.93, 95%CI: 1.11-3.41).

Late adopters and those who did not receive any SMC reported significantly longer travel time to the nearest health facility. In contrast, those who dropped out before finishing SMC lived closer to a health facility (Table 3).

**Table 3:** Summary of SMC coverage, mean time to travel to a health facility, and knowledge of community health services in SMC coverage categories.

|  |  |  | *Time to health*<br>*facility (min)* | *Know CHP* |
| --- | --- | --- | --- | --- |
|  | *N* | *Mean, 95% CI* | *Mean, 95%CI* | *Mean, 95%CI* |
| Full SMC | 479 | 71.7%<br>(64.8-78.7) | 71.1<br>(42.4-99.7) | 99%<br>(98-100) |
| Late adopter | 57 | 8.3%<br>(4.9-11.8) | 97.2<br>(39.2-155.3) | 79%<br>(56-100) |
| Dropped out | 52 | 6.2%<br>(3.1-9.3) | 31.9<br>(18.9-44.9) | 97%<br>(92-100) |
| Partial (Other) | 60 | 10.3%<br>(6.8-13.7) | 47.8<br>(26.8-68.9) | 100%<br>- |
| No SMC | 23 | 3.5%<br>(0.8-6.1) | 109.4<br>(7.3-211.5) | 73%<br>(34-100) |
| Total | 680 | 100% | 67.5<br>(43.9-91.0) | 96%<br>(92-100) |

### All estimates are adjusted for the sampling design

Similarly, late adopters and those without SMC were less likely to know their CHP. To parse factors differentially associated with dropout and late adoption, we compared models with late adoption or dropout as the outcome to other partial SMC (S1 Table, S2 Table). We noted that children whose family knew the CHP had lower odds of late adoption (Adj OR=0.04, 95%CI:0- 0.25), but children who lived furthest from the facility had higher odds of joining SMC late (Adj OR=2.95, 95% CI: 0.93-9.55). Although wealth was not a factor in late adoption, the least poor families were three times *more* likely to drop out (Adj OR=4.61, 95% CI: 1.53-15.48). Other factors were not significantly correlated with these categories of partial SMC uptake.

The qualitative results provide further insight into the factors influencing adoption. First, caregivers across all categories (non-adopters, partial adopters and full adopters) perceived malaria to be a serious illness and a threat to their family. This was mainly due to their personal and/or second-hand experiences with the severity of malaria. A considerable number of caregivers across the three categories revealed having known someone who died as a result of malaria. Caregivers highlighted that children are not only more susceptible to malaria but also experience its effects more severely compared to adults. Ambivalence about malaria was not a factor in the poor adoption rate.

The quantitative analysis of factors related to complete SMC uptake strongly implicated the role of the CHP. This was reinforced in interviews with full and partial adopters. The CHPs, who were the largest distributors of SMC, had a significant influence on caregivers’ uptake of SMC. Caregivers’ assessment of distributor credibility was associated with three attributes – familiarity, trust, and perceived knowledgeability. Familiarity stemmed from long-standing social ties. The caregivers stated that they knew the CHP (very) well as they had known each other since childhood, resided in the same locality/village, the CHPs were assigned to the caregiver’s locality, and/or they knew the CHP were stationed at the local health facility where the caregiver sought care.

> *I know her very well; she is from here. **(SMC_FA_11)***

Conversely, there were specific instances in which children missed out on SMC due to the drug distributors’ unfamiliarity with the community, as they were not the usual CHPs assigned to the area.

> ***Interviewer:*** *Why did the child not get SMC medicine at that time?*

> ***Respondent:*** *I don’t know where the problem was. It is like we didn’t cross paths again with those SMC distributors. However, since [name of usual CHP] began to deliver the SMC medicine, my child never missed because he brought it to her cycle, that is, for the September and October cycles. **[SMC_PA_06.]***

Beyond familiarity, the caregivers trusted the CHPs who offered them SMC. This trust was attributed to the aforementioned long-standing acquaintanceship and to the CHP’s professional position, which entails working closely with health facilities on health issues, especially children’s health. Partial adopters further stated that they trusted the CHPs who offered them SMC because they were externally validated: these CHPs had received training on SMC delivery; competent health entities verified the products and/or information that they were offering; relevant authorities had identified these CHPs as the personnel fit to distribute SMC; and the community had been informed of the CHPs role in SMC distribution during the public sensitization period. In some instances, mistrust of the CHP, arising from cited bad behavior or lack of familiarity, led to SMC refusal.

> ***Interviewer:*** *Do you think the CHPs were knowledgeable on matters SMC?*

> ***Respondent:*** *Yes, because he has been taking care of our children, so I trusted him, and I believed in this new adventure, he must have been trained. **(SMC_FA_12)***

> *I trusted him because he has always taken care of our children, and I know him, he is from here. **(SMC_PA_01)***

> ***Interviewer:*** *You declined because you did not know her?*

> ***Respondent:*** *I declined because she was drunk. **[SMC_PA_03]***.

The final credibility attribute was the perception of the distributor’s knowledge regarding malaria and SMC. Some respondents had *a priori* confidence in CHPs’ knowledgeability based on established trust and experience, whereas some built confidence in the CHPs’ expertise through the SMC interaction. Caregivers expressed satisfaction with the comprehensive information received from the drug distributor, including answering any questions they had and addressing their concerns [laying to rest any fears they had]. Full adopters further perceived SMC distributors as knowledgeable because, when they (caregivers) followed the SMC distributor’s instructions for administering SMC, the children did not experience any side effects, and the information CHPs provided on SMC’s primary function was validated, as the children did not contract malaria after taking SMC.

> ***Respondent:*** *I believed her because after that, there was a time my child had a fever, and I thought it was malaria. I almost doubted if the [SMC] worked, but when I took him to hospital they tested and said he does not have malaria. It was just the flu…*

> ***Interviewer:*** *Okay. So, the explanation that the CHP gave you was correct?*

> ***Respondent:*** *Yes, I realized that it was correct **(SMC_FA_01)***

Although the overall picture shows CHPs positively influencing uptake of SMC, several respondents cited negative attitudes of other healthcare workers towards SMC. This discouraged their participation and was noted as one of the reasons caregivers would decline future participation in SMC.

> *And also, the way some service providers were talking about [SMC], it seems that they were not for it. They are against [SMC]. **…** And then the health service providers are the ones who talk negatively about [it], what can ordinary people do in this case? **(SMC_NA_12)***

The factor cited most often in explaining sustained uptake of SMC over the course of the transmission season was experience with SMC’s effectiveness in preventing malaria. Full adopters commented that they had observed reduced malaria cases among their own children and in the broader community, which solidified their confidence in the effectiveness of SMC. Partial adopters were more likely to cite the lack of side effects in their children as a reason for continuing.

> *Seasonally, we normally experience serious malaria that we perceive as quite deadly, but we seek medical care. However, recently we’ve not had such a serious case especially after the SMC drugs were brought. **(SMC_PA_13)***

> *There was no negative outcome. The child was okay, he never had any adverse effects, he never got sick throughout the SMC period, he only caught the flu. **(SMC_PA_13)***

Additional motivators that partial and full adopters cited as facilitating their participation were: protecting the children from an impending/ongoing outbreak of malaria; avoiding future malaria- related treatment costs; perceiving SMC as safe for the children; and public sensitisation that resulted in caregivers’ sound understanding of SMC. They further cited their confidence in SMC as a key motivator to allow their children to take up SMC. Their confidence was based on SMC being a product approved by trusted authorities, specifically the government and health facilities; SMC being provided to all children in the community; and community members allowing their children to take up SMC.

> ***Respondent:*** *I was expecting it to help because the government has good plans for the children*.

> ***Interviewer:*** *How did you know it would help the child?*

> ***Respondent:*** *I didn’t. I trusted what the CHP said and the government*.

> ***Interviewer:*** *Were you worried about it at some point?*

> ***Respondent:*** *No. because it was intended for all children in the community*.

> ***(SMC_PA_10)***

Additionally, partial adopters revealed that being provided with SMC at no extra cost motivated them to take up SMC. Their confidence in SMC’s reliability encouraged uptake to avoid future malaria-related treatment costs.

> *… the medication was free. **(SMC_PA_01)***

### Factors related to the late adoption of SMC

Two key understandings emerged from the evaluation of late adopters’ responses. First, several late adopters were forced to wait for their child to join SMC because of illness at the beginning of the SMC program. Caregivers reported that SMC distributors advised against enrolling children who were unwell and undiagnosed, unwell and on treatment, and/or just finished treatment for any other illness, especially malaria. With SMC being initiated after the beginning of the transmission season, it was not surprising that some children had been diagnosed with malaria and were on treatment or had recently completed their malaria treatment. Drug distributors were unable to perform malaria point-of-care testing at home. They were therefore required to refer any children who were unwell on the day of distribution to the health facility for testing for malaria.

> ***Respondent****: I was only talking about it with that CHP. When she explained to me [about SMC], I told her that my child had just been on malaria treatment. I asked her what I should do. She told me to wait until that cycle is complete, then she will give it to him in the next cycle. **[SMC_PA_02]***

The second reason cited by late adopters for their children’s delayed enrollment in SMC was the caregiver’s uncertainty about SMC. They revealed that being new raised concerns about its safety and efficacy. Sometimes these concerns were influenced by negative opinions from people in close relationships with the caregiver, especially family members. For some full adopters, and particularly among partial adopters, the decision to accept SMC for their child was shared with the spouse/partner, influenced by neighbours and other community members, and arrived at after consultation. Often, they expressed a ’wait and see’ approach:

> *The first time the other children were offered SMC, he did not receive it. But when I saw that the other children who had received it had not experienced any negative effects, they were just fine, I thought I should also allow my child to also receive it (referring to SMC). "Let me not be left behind, this medication could actually be good." So, I also released my child. I permitted my child to be given SMC. I accepted it during cycle 3. **[SMC_PA_09]***

> *We observed from others who were given and then decided to give him later. Then we agreed for him to get it because the ones who were given first had no side effects. So, we knew, yes, it is good. Let ours also be given. Yes. **(SMC_PA_09)***

Safety concerns were further fueled by long-standing rumors in the community about a previous polio vaccine campaign that resulted in adverse health outcomes for all the children who received this vaccine. Therefore, for this category of caregivers, they thought it wise to wait and observe how other children in the community who received SMC would respond to it before permitting their children to take it up.

> *I know some people…a long time ago we would hear children got polio vaccines. I do not know which polio vaccine, but they all became ruined later. "So, if this is new, let us observe it from others first." That is what almost caused difficulty. Yes. **[SMC_PA_09]***

### Non-adopters

Caregivers whose children did not receive any SMC shared some of the concerns cited by partial adopters. A few reported that during the SMC distribution period, their child either had an already known underlying medical condition, was generally in poor health, presented as a sickly child, or was sick and receiving treatment at every cycle. The SMC distributors excluded these children from SMC. Some caregivers, especially those whose children were ill, agreed with the distributors’ decision to exempt the child out of fear that uptake of SMC would place the child at a higher risk of experiencing adverse side effects. Other caregivers expressed their willingness to participate in SMC if their children met the eligibility criteria.

> *I know it could have helped my child, but according to the information that we were being given, we were told that if the child is sick or is sickly, that child should not get the [medication]. So, because mine is a sickly child who has had problems with the umbilical cord, tonsils, gets sinuses and I take her to hospital frequently, I thought that maybe the [medication] might affect her badly. That is why I decided not to let her get [SMC]. **[SMC_NA_03]***

> *Because it helps by preventing children from getting malaria. Even at that time, it is just that my child was sick throughout until the period ended and therefore, he missed on getting the drugs. That is why he did not get it. He was sick during that period. The time he got well, and I thought I would manage to get it for him, the period of the program was already over. The child was sick all the time **[SMC_NA_08]***

More often, it was the caregiver’s skepticism about SMC. This skepticism stemmed primarily from the recent introduction of SMC in the local community. Consequently, it led to doubts about SMC’s effectiveness in preventing malaria, concerns regarding SMC’s safety, and concerns regarding the availability and accessibility of quality healthcare services in case a child experienced SMC side effects. Some safety concerns were built on the perception that SMC was an experimental intervention being tested for the first time in Turkana Central. A few caregivers chose not to share their concerns with the SMC distributors for undisclosed reasons. They instead accepted the SMC medicine from the distributors but opted not to administer it to the child(ren).

> *I was wondering what would happen to my child if he is given that [drug]. I was wondering if it will harm my child or if the child will be okay. That is what I was thinking about **(SMC_NA_01)***

> *Because I heard some people saying that they have come to test the performance of [SMC]. That is why they fear giving it to their children. **(SMC_NA_12)***

> *I asked them that in case the drug reacts on my child, and I take the child to the district hospital, will the child get good treatment there? **(SMC_NA_01)***

> *The concerns I had were very personal, I didn’t want to tell the CHP I didn’t want the medication, but I took it. I was skeptical about it. **(SMC_NA_11)***

While uncertainty about SMC led some caregivers to reject it outright, others chose to postpone enrolling their children, opting instead to observe how other children in the community responded to SMC before making their decisions. However, community members’ negative experiences with SMC discouraged them even further after witnessing or hearing reports of previously healthy children becoming ill after receiving SMC, with some children suffering moderate to severe side effects that necessitated medical attention at a health facility. Moreover, some caregivers who delayed enrolling their children expressed doubts about the effectiveness of SMC, questioning whether it would function as intended if a child skipped earlier cycles and was only enrolled in the later stages of the program. Ultimately, these caregivers did not enrol their children in the program.

The non-adopters disclosed additional reasons for not enrolling their children in SMC, exclusive to this category of caregivers. One such reason was being missed out by SMC distributors, either because caregivers lacked awareness of SMC and distribution dates or because some families lived in remote areas.

> **Respondent:** I was not aware of the medication exercise. **(SMC_NA_05)**

Another reason for non-enrollment was perceiving SMC as unnecessary because their child had never contracted malaria, or that treatment was readily available, so prevention was unnecessary.

> *The way it was publicized, they said it prevents malaria. So, I thought that because this child has never suffered malaria, there was no reason to give him. He normally just gets colds, but he has never gotten malaria. **(SMC_NA_02)***

> ***Interviewer:*** *So you will not participate even next time?*

> ***Respondent:*** *Yes, I will not participate*.

> *Interviewer: Why?*

> ***Respondent:*** *When they contract malaria, they will just get treated. (**SMC_NA_04)***

## Discussion

This study demonstrates that high seasonal malaria chemoprevention (SMC) coverage is attainable in Turkana County, a large, arid region in northern Kenya characterised by highly seasonal malaria transmission and mobile pastoralist communities. The main finding that 97% of eligible children aged 3 to 59 months received at least one cycle of SMC, with a complete coverage rate of 71% across five cycles, is a significant achievement for the first SMC program in Kenya. This coverage is comparable to that reported in the Sahel region [10], where SMC has been in use for over a decade, indicating that adapting SMC for mobile populations is both feasible and highly effective [11,12].

The program’s success can be attributed to its mixed-model distribution approach. By combining door-to-door drug distribution through CHPs with mobile fixed points and integrated outreach efforts, the program effectively addressed the distance barrier in the Turkana Central area, where travel time to the nearest health facility is often more than an hour. The absence of a relationship between time to travel to a facility and SMC coverage in our study demonstrates the effectiveness of the adapted community-based delivery strategy in overcoming disparities in access to health facilities. Furthermore, the program achieved high adherence to subsequent doses, with 95% reported adherence on day 2 and day 3. Community acceptance was also notable, as only 8.7% of non-recipients expressed concerns about safety or benefits. These factors highlight the acceptability and potential sustainability of SMC in this community.

Door-to-door distribution has been shown to increase SMC coverage in the Sahel. A study in Nigeria found that caregivers reported it was convenient because it did not disrupt their daily tasks [13]. Combined approaches to complement door-to-door delivery may be more suitable for nomadic populations and hard-to-reach areas. Fixed points were found to add value to SMC delivery in Turkana, particularly “mobile fixed points” in a place chosen by the community (such as a school or a main meeting place under a tree), and fixed points organised as outreaches combined with other health services. In a single village, we found that 85% of children received SMC through a mobile fixed point. Mobilizing caregivers in remote areas to assemble at a fixed point reduced challenges for drug distributors covering vast areas and, more importantly for this region, reduced challenges with finding households that have migrated.

Drug reactions were rare in our study population, consistent with SMC programs in the Sahel [14]. Although having a reaction was not statistically associated with dropping out or receiving partial SMC, several caregivers reported that their experience with drug reactions was a barrier to SMC uptake in subsequent months, especially the fear of not knowing what action to take in case of drug reactions, which affected SMC uptake in this population. Only 77% of caregivers reported receiving information about side effects from the drug distributor. Strengthening this aspect of CHP training and household communication could reduce fear and improve acceptability.

Several factors contributed to differential SMC adoption among families. Knowing the CHP in their village and discussing SMC with others were strongly correlated with completing SMC. At the first level, several villages lacked a CHP and were missed in the initial cycles of SMC because the health system had not mapped them. These villages tended to be remote, as evidenced by late adopters having significantly longer travel times to a health facility. In addition, CHPs are trusted health ambassadors and often take up health delivery roles in many countries [15], including as drug distributors for SMC [16]. Their knowledge and ability to communicate effectively with households enhanced the acceptability of SMC in Turkana. CHP training on key aspects of SMC is critical to their credibility and ability to transfer knowledge to target populations.

On the other hand, our data also reveal that the lack of participation of other healthcare providers was a barrier to SMC uptake. This calls for advocating SMC among all health workers to support and participate in SMC, as their lack of participation affects community decisions on SMC uptake. Notably, delayed participation in SMC amongst some children was attributed to a child having malaria during the first month of SMC or recently completed their treatment and were therefore excluded from SMC. This emphasizes proper timing of the SMC programme to ensure children are protected before the peak transmission season. Wealthier households were more likely to receive partial SMC and especially to drop out of the programme early, possibly due to economic activities which took caregivers away from home during the day, or to feeling confident in their ability to access care if the child contracted malaria thus reducing the perceived importance of prevention, or to the bias against receiving health services from lay health workers (CHPs).

In the cross-sectional design, households that were randomly selected to participate but were unavailable on the day of the survey and were still away on the day of the mop-up visit were replaced with households where the caregiver was present. This may have underestimated the effect of the nomadic lifestyle on coverage. However, we attempted to maintain the representativeness of the sample by randomly pre-selecting replacement households with the same probability as the primary sample. We also relied heavily on CHP and village elder knowledge about the community to generate household lists. Our study demonstrated that households that did not know their CHP were much more likely to be missed, especially in the early cycles of SMC. This same effect could have biased our sampling frame. Finally, we often had to rely on caregiver reports for the number of SMC cycles a child received. It is possible that recall was less accurate for cycles that occurred months before the survey. This could have inflated the observed coverage. We tried to mitigate this by speaking with the caregiver of each child if more than one caregiver was living in that household, by comparing the caregiver report to the physical record (SMC card) whenever possible, and by grouping partial SMC recipients in our analysis rather than relying on the exact number of cycles as our outcome.

This study reveals that SMC is a feasible intervention in areas with semi-nomadic populations with limited access to health infrastructure, and that the household-focused programme can reduce disparities in access that plague health interventions in this remote area. SMC coverage was high and improved with each cycle as the delivery model evolved to meet the unique needs of the communities. Tailoring drug distribution strategies in areas with limited infrastructure and semi-nomadic populations is critical to ensuring no child is missed. The quality of the CHP interaction at the household level resulted in high acceptability and adherence to the 3-day regimen. The caregiver’s understanding of the SMC programme, including expected side effects, was a critical factor in SMC acceptability and continuity. The high coverage, adherence, acceptability, and willingness to participate in future SMC campaigns indicate the need for urgent scale-up.

## Data Availability

The data will be made available in a public repository after acceptance of this manuscript.

## Acknowledgements

We are grateful to the study participants without whom the study would not have been conducted. We also acknowledge the research assistants: Patrick Ejore, Zubeda Ewoi, Chuchu Gregory, Joseph Lobee, Lilian Lokitoe, Emmanuel Lopeto, Kaman Lowoi, Purity Musai, Joyce Nachipon, and Jeremiah Nokorot. We appreciate the contributions of Caroline Ngina, Tabitha Chepkwony, George Ambani and Emily Robie. The partnership and leadership of the Turkana County Health Team was key to the success of this work.

## Author statement

The authors declare no conflict of interests.

